# A Voice App Design for Heart Failure Self-Management: A Pilot Study

**DOI:** 10.1101/2022.04.06.22273509

**Authors:** Antonia Barbaric, Cosmin Munteanu, Heather J. Ross, Joseph A. Cafazzo

## Abstract

There is a growing interest to investigate the feasibility of using voice user interfaces as a platform for digital therapeutics in chronic disease management. While mostly deployed as smartphone applications, some demographics struggle when using touch screens and often cannot complete tasks independently. This research aimed to evaluate how heart failure patients interacted with a voice app version of an already existing digital therapeutic, *Medly*, using a mixed-methods concurrent triangulation approach. The objective was to determine the acceptability and feasibility of the voice app by better understanding who this platform is be best suited for. Quantitative data included engagement levels and accuracy rates. Participants (n=20) used the voice app over a four week period and completed questionnaires and semi-structured interviews relating to acceptability, ease of use, and workload. The average engagement level was 73%, with a 14% decline between week one and four. The difference in engagement levels between the oldest and youngest demographic was the most significant, 84% and 43% respectively. The *Medly* voice app had an overall accuracy rate of 97.8% and was successful in sending data to the clinic. Users were accepting of the technology (ranking it in the 80^th^ percentile) and felt it did not require a lot of work (2.1 on a 7-point Likert scale). However, 13% of users were less inclined to use the voice app at the end of the study. The following themes and subthemes emerged: (1) feasibility of clinical integration: user adaptation to voice app’s conversational style, device unreliability, and (2) voice app acceptability: good device integration within household, users blamed themselves for voice app problems, and voice app missing desirable user features. The voice app proved to be most beneficial to those who: are older, have flexible schedules, are confident with using technology, and are experiencing other medical conditions.

## Introduction

### Background

Chronic diseases are the leading cause of death and disability worldwide, with over 41 million people dying each year due to these diseases (1). Cardiovascular diseases, such as heart attacks and high blood pressure are responsible for most chronic disease deaths (17.9 million people) (1). Patient self-care is considered to be essential in the prevention and management of chronic diseases (2) as studies have shown the benefits of this approach, which include improved health outcomes, decreased clinic visits, and decreased health costs (3). Mobile health, also referred to as mHealth, is a type of digital health technology that involves the use of mobile devices for medical and public health practice (4), and enables the integration of self-care support into a patient’s routine (5). While mHealth apps are one of the most popular tools for helping patients with chronic conditions manage their health at home (6), there is research to suggest voice apps are an emerging platform that will create alternative interaction models that some patient demographics may find more accessible.

Voice user interfaces (VUIs) are becoming more prevalent in the healthcare field for a variety of different purposes. With VUIs the user is able to interact with a computing system using only speech, with voice apps being one example of this technology. The primary advantage of implementing VUIs in any environment is simplicity, since it does not require the user to interact with a hand-held technology, as we are typically accustomed to. So far, VUIs have been used to help those who have speech or hearing difficulty, to improve patient engagement, as well as aging in place (7). This technology has also been used in the clinical setting by supporting physician note transcription and the patient registration process (7). Devices that offer VUI capability are also continuing to gain popularity in consumer households and are becoming more integrated into our daily lifestyles due to their convenience, ease of use, and affordability. As a result, there is growing interest to investigate the feasibility of using smart speakers to improve patient engagement, with a specific focus on chronic disease management.

### Heart Failure

Previous research has begun to investigate the potential for patients to manage their heart failure (HF) using a voice app (publication pending). HF is a condition that develops after the heart muscle becomes damaged or weak due to cardiovascular diseases, such as heart attacks and high blood pressure. When the heart muscle becomes damaged or weakens, it is unable to pump enough blood to meet the body’s needs for blood and oxygen (8). *Medly* is an evidence-based, HF self-management program that has been developed by the University Health Network (UHN) and is implemented as part of the standard of care at UHN’s Ted Rogers Center of Excellence for Heart Failure clinic (9). This program is currently available to patients through a smartphone app and enables them to log clinically relevant physiological measurements and symptoms, which is then used in the *Medly* algorithm to generate an automated self-care message. A voice app version of *Medly* has already been built as part of previous work, and a usability study has been performed with the voice app at UHN’s Heart Failure Clinic.

### Objectives

The results from a previous usability study show promise that a voice app for chronic disease management, such as HF, is feasible to deploy and acceptable by patients (publication forthcoming). With these findings, we sought to perform a more in-depth clinical evaluation by using *Medly* as a case study, with 20 HF participants. The goal of this study is to determine if voice apps can be a practical alternative of enabling patients to receive a digital therapeutic. The following research question will be investigated:

*What is the acceptability of a voice application for patients, through the use of a smart speaker, for a home chronic disease management platform?*

Through this research we hope to uncover whether deploying a voice app version of *Medly* adds any benefit to the current model of interaction and care.

## Methods

### Participant Recruitment

HF patients were asked to participate in this study and interact with the *Medly* voice app while in their homes for a four week period. Participants were considered eligible if they had been diagnosed with HF by a physician at UHN’s HF clinic, and were prescribed the *Medly* program. Participants were also required to speak and read English adequately to understand the voice prompts in the *Medly* application. The *Medly* nurse coordinator first provided a brief overview of the research study to interested patients prior to introducing them to the study coordinator. If they agreed to participate, informed written consent was obtained by the study coordinator prior to onboarding.

A total of 20 participants were recruited for this study, based on findings from the literature which suggested that a sample size between 10 and 30 users (10) is appropriate to use for pilot studies. To help mitigate potential bias, we aimed to recruit both participants who were ‘new’ to *Medly* (less than 2 month since being onboarded to the program), and also those who were ‘existing’ (more than 2 months since being onboarded) *Medly* patients. In the end we recruited a total of 7 new and 13 existing *Medly* patients.

Recruited participants were required to perform a double-entry of their *Medly* measurements for the four week duration, more specifically they were asked to first input their *Medly* measurements on the smartphone app prior to interacting with the voice app. Each participant received a gift card to compensate them for their time participating in the study. Ethics approval was obtained from the University Health Network (UHN) Research Ethics Board (20-6095).

### Study Outcome Measures

The evaluation of the *Medly* voice app was influenced by two frameworks: Proctor et al.’s Implementation Outcomes (11) and Unified Theory of Acceptance and Use of Technology 2 (UTAUT2) (12). The outcomes listed below were selected from both frameworks to help guide the quantitative and qualitative data that was gathered to determine the acceptability of the *Medly* voice app.

Questionnaires and semi-structured interview questions were influenced by the System Usability Scale (SUS) (13) and NASA Task Load Index (TLX) (14) standardized assessment tools. Quantitative data was also gathered through semi-structured interviews by asking participants how often the voice app misheard their measurements, how many times they were required to correct wrongly recorded data, and how many times they missed inputting their measurements and why (engagement levels). Accuracy rates were calculated by comparing the measurements inputted on the smartphone app versus the voice app.

### Data Collection

The study coordinator performed an onboarding session over the phone with each participant to help them set-up and access the *Medly* voice app, and provided them with an instructions manual (Fig S1, Multimedia Appendix 1). Participants were also required to answer a baseline questionnaire (Table S1, Multimedia Appendix 2) to help the study coordinator understand their comfort levels with using technology. Participants were made aware that they needed to perform a double entry of their *Medly* measurements for the four week duration and were told to prioritize the *Medly* smartphone app, namely to input measurements on the phone first, and to only follow guidance from the smartphone app.

A one week check-in was scheduled with all participants over the phone. During this check-in, the study coordinator asked each participant about their experience so far and collected the quantitative data described above. Following the week one check-in, participants were emailed a questionnaire at the end of week two. The questions were influenced by the frameworks and standardized assessment tools mentioned earlier in order to gauge the participant’s thoughts and opinions of using the voice app thus far. As part of each participant’s offboarding, the questionnaire was sent out again (to see if thoughts or opinions changed), and a semi-structured interview took place with the study coordinator over the phone.

### Study Analysis

Descriptive statistics for the standardized questionnaire responses were performed and recorded using Microsoft Excel. Graphical representations of engagement levels were also created using Excel. The responses from the System Usability Scale questionnaire were analyzed as per standard protocol (13) and averages were calculated for the NASA-TLX and UTAUT2 questionnaires, both overall and question specific. Data was categorized in different ways using various characteristics and then analyzed to identify any trends or commonalties.

The quantitative data was then triangulated with the qualitative data findings, namely from the semi-structured interviews. Interview transcripts were analyzed by the study coordinator (AB) using an deductive approach. Specifically, the transcripts were analyzed under the guidance of Proctor et al.’s Implementation Outcomes framework, with a focus on the acceptability and feasibility constructs. Sub-themes were then identified to better describe the study findings. The transcripts and coding was organized using Microsoft Word.

## Results

### Characteristics of Study Participants

A total of 20 patients were recruited for the study, with a fairly even split among genders (females: 9/20, 45%, male: 11/20, 55%) and an average age of 57.8 (SD 13.1) years. All patients who were recruited were required to be enrolled in the *Medly* program, with a mix between those who have just recently (defined as less than 2 months since the time of recruitment) been onboarded to the program (7/20 patients, 35%) and those who have been enrolled in the program for longer (13/20 patients, 65%). Other patient characteristics were also collected for the purposes of this study, such as: comfort levels with technology and whether or not they have used a smart speaker before through a baseline questionnaire (18 out of the 20 participants returned this questionnaire). The statistics for each of these characteristics can be seen in Table 2.

**Table 1.**
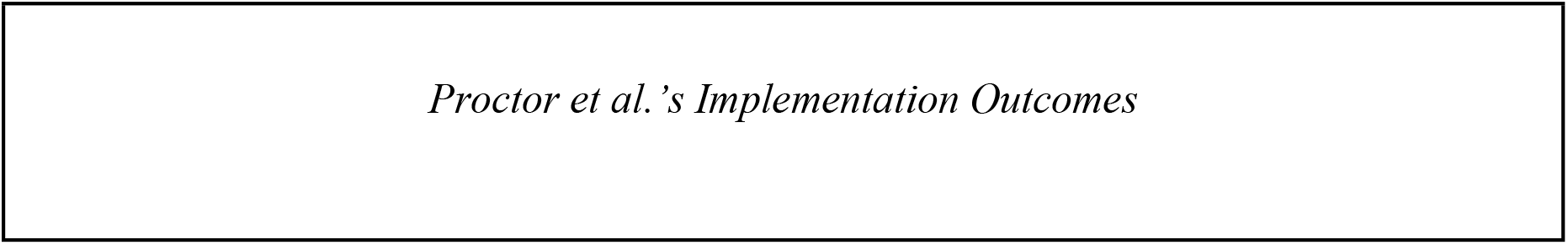
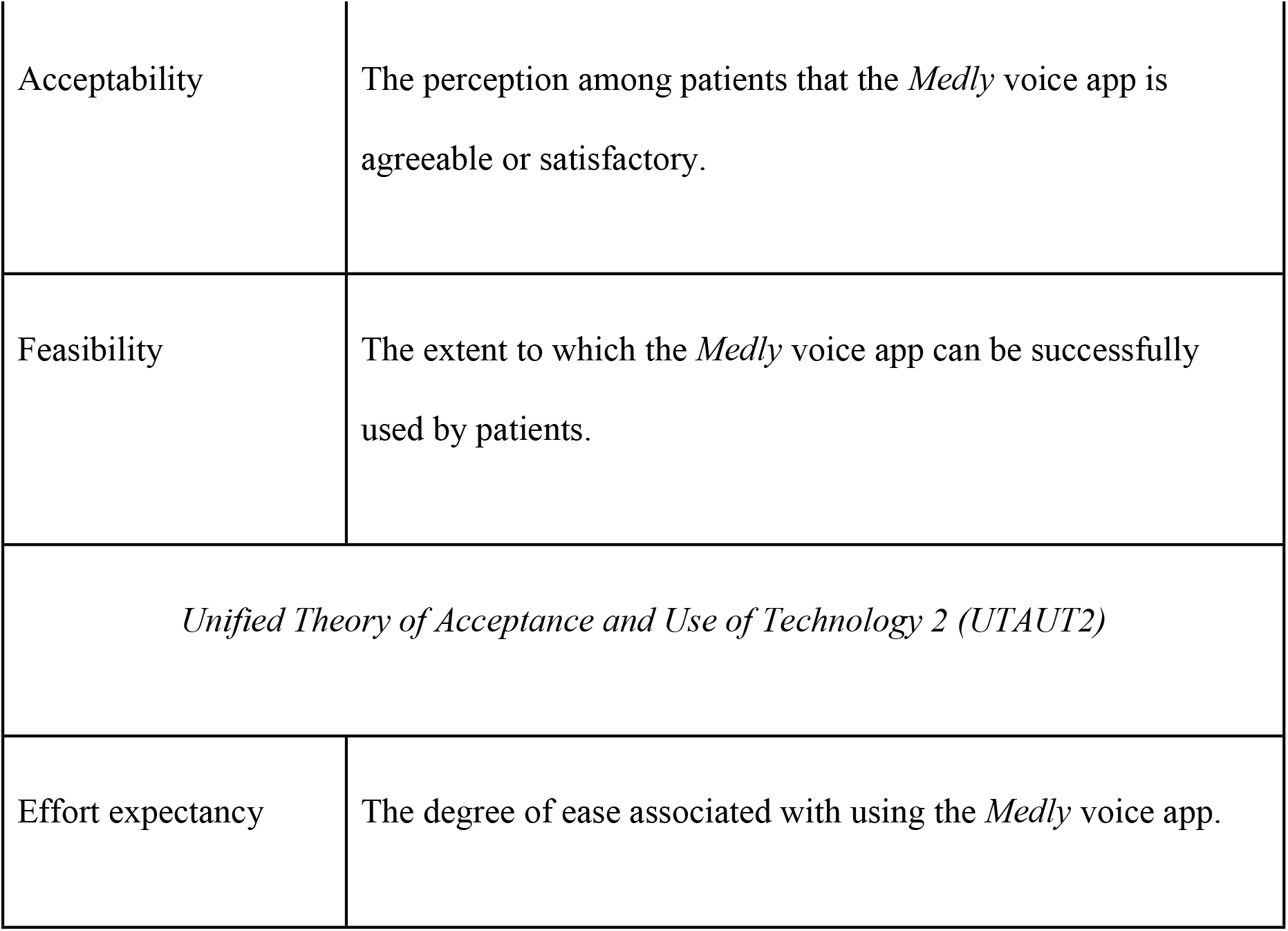
Definitions of the selected outcomes chosen from frameworks to identify the acceptability of using the *Medly* voice app.

|  |
| --- |
| *Proctor et al.’s Implementation Outcomes* |

|  |  |
| --- | --- |
| Acceptability | The perception among patients that the *Medly* voice app is agreeable or satisfactory. |
| Feasibility | The extent to which the *Medly* voice app can be successfully used by patients. |
| *Unified Theory of Acceptance and Use of Technology 2 (UTAUT2)* |  |
| Effort expectancy | The degree of ease associated with using the *Medly* voice app. |

**Table 2.** Patient characteristics used to categorize and sort data in the study.

| **Characteristics** | **Statistics** |
| --- | --- |
| Age (years), mean (SD) | 57.8 (13.1) |
| **Age (years; categorical), n (%)** |  |
| 20 or younger | 0 (0.0) |
| 21 - 40 | 2 (10.0) |
| 41 - 60 | 7 (35.0) |
| 61 - 80 | 11 (55.0) |
| 81 or older | 0 (0.0) |
| **Sex, n (%)** |  |
| Female | 7 (35.0) |
| Male | 13 (55.0) |
| **Interacted with smart speaker before, n (%)** |  |
| Yes | 7 (35.0) |
| No | 11 (55.0) |
| **Comfort level with technology, n(%)** |  |
| Very uncomfortable | 1 (5.0) |
| Somewhat uncomfortable | 0 (0.0) |
| Neutral | 6 (30.0) |
| Somewhat comfortable | 2 (10.0) |
| Very comfortable | 9 (45.0) |

## Quantitative Data

### Engagement Levels and Accuracy Rates

The overall engagement level for the entire study population during the four week period was 73%, with noticeable drops in engagement as the weeks progressed (Table 3) and an overall decline of 14% when comparing week one and four averages.

**Table 3.** Overall and weekly average engagement levels over the four week study duration.

|  |  |  |  |
| --- | --- | --- | --- |
| Overall engagement level/SD (%) |  | 73.4 / 9.5 |  |
| *Week #* | *Average (%)* | *Average Days Missed* | *SD (%)* |
| 1 | 80.7 | 1.4 | 11.3 |
| 2 | 75.0 | 1.8 | 5.8 |
| 3 | 70.7 | 2.0 | 7.9 |
| 4 | 67.1 | 2.3 | 8.1 |

In addition to calculating the overall engagement levels, patient characteristics from Table 3 were also used to group the study population and compare the results among sub-groups to identify any noticeable trends. These results can be seen in Table S1, Multimedia Appendix 3.

In summary, there were no noticeable trends when comparing voice app engagement levels among new and existing *Medly* patients, with an ∼1% difference overall. Similar to the findings related to the entire study population, engagement levels were lower in week four when compared to week one for both of these groups.

In contrast, engagement levels when compared to the different age groups showcased more obvious trends. Overall, average engagement levels increased as the age groups increased, with the oldest demographic (61-80 year olds) having the best engagement level of 84.1%, almost double the overall engagement level when compared to the youngest age group in the study. Those in the 61-80 age group were also the most consistent throughout the four week duration and had the smallest difference between the different engagement week averages.

A similar trend was observed when comparing participants based on their reported comfort levels with technology. Those who were very confident consistently used the technology more through the four weeks than those who reported less confidence (13.6% overall difference). There were also consistently higher engagement levels with the group that had never interacted with smart speakers before when compared to those who have (7.6% difference). Both groups steadily declined in engagement as the weeks progressed, with similar overall differences between week one and four averages.

Over the four week duration (28 days), nine entries (out of 411) were incorrect measurements that were submitted using the *Medly* voice app, indicating an overall accuracy rate of 97.8%. The errors varied between weight and blood pressure. A small subset of participants (four) were not able to successfully submit their correct readings which led to the nine errors that were recorded.

### Acceptability of the *Medly* Voice App

Findings from the SUS questionnaire paired with the findings from the semi-structured interviews were used to better understand the acceptability of using a voice app version of the *Medly* program.

The responses from the SUS questionnaire from week two resulted in an overall average score of 69 (out of 100), ranking it in the 53rd percentile based on previous studies. In contrast, the average score from week four results is 77 (out of 100), ranking it in the 80th percentile based on previous studies. This data indicates an overall increase in the level of satisfaction of using the *Medly* voice app (by 27%) from the study population. The difference in averages for each individual question between weeks two and four was also calculated, with the last question in the survey having the biggest difference of 13%. Participants felt that as time went on they needed to learn more things about the voice app to successfully interact with it (consistent with NASA TLX cognitive load results). Response distributions for week two and four results were fairly similar for all of the questions (Fig S1, Multimedia Appendix 4).

Average SUS scores were also calculated based on the different patient characteristics (age, *Medly* status, comfort levels, and interaction with smart speaker familiarity). Overall the scores were similar in range for all of the characteristics. The largest range in data however, was identified in the age groups, with the oldest (61-80 years) demographic providing the lowest score (72 out 100), ranking it in the 62nd percentile, while the middle-age demographic provided an average score of 87.5 out of 100, ranking it in the 96th percentile. The average score from the youngest demographic was 77.5, ranking it in the 80th percentile.

### Feasibility of *Medly* Voice

The NASA-TLX was used in this study to better assess the perceived workload when using the *Medly* voice app by the study participants. A 4% increase was seen in average scores between week two and four results, indicating a slightly higher workload. While the averages for each of the questions were fairly low, questions relating to: 1) success rates, 2) how hard they needed to work to accomplish the task, and 3) feeling of discouragement, irritation and stress scored the worse when compared to the rest of the questions. These results can be seen in Fig S2, Multimedia Appendix 4. Participants also felt less successful with using the *Medly* voice app at the end of the study than they did at the end of week two (22% difference in results).

When analyzing the scores based on the different age groups, it was found that the youngest demographic felt they needed to work the most (highest average of 2.67) when compared to the middle-age (average of 1.61) and oldest demographics (average of 2.12). It was also found that those who were new to *Medly* specifically felt more rushed when using the voice app and less successful when inputting their measurements, when compared to those who have been on the *Medly* program for a longer time (∼15% difference in scores for each question). The difference in average scores for those who described themselves as less confident when using technology consistently gave poorer scores for each of the questions, indicating they had a more difficult time than those who described themselves as confident (Table S1, Multimedia Appendix 4).

In summary, the youngest age group felt they needed to work the most, the study population collectively felt they needed to provide slightly more effort as time went on, and those who were less familiar with technology had more difficulty using the voice app when compared to those who were more confident.

The UTAUT2 questionnaire was used to better understand participants’ thoughts regarding facilitating conditions, effort expectancy, habit, and behavioral intention when it comes to using the voice app. The biggest difference between week two and four results was with whether they would use the *Medly* voice app in the future, with a 13% decline in the average score. The oldest demographic was the least keen on using it in the future, while the middle-aged demographic was the most interested in future use. When asked if the voice app became a habit, those who used the technology before agreed more than those who did not have experience using the device (19% difference in responses).

Overall, all participants felt the voice app required low effort to use, and that it was easy for them to operate. They were less certain with whether or not using the voice app had become a habit for them (this can be supported with engagement levels), and were least certain about whether they would use the voice app in the future, as seen in Table S2, Multimedia Appendix 4.

## Qualitative Data

Interview themes were classified using Proctor et al.’s Implementation Outcomes, specifically focusing on the *feasibility* and *acceptability* constructs to better understand which patient demographics would benefit from using the voice app. The themes: (1) feasibility of clinical integration and (2) voice app acceptability are presented below, each with their own set of accompanying sub-themes.

### Feasibility of Clinical Integration

The feasibility of providing patients with the option of accessing DTx through voice apps in the clinic first involves understanding how patients interact with the technology, as well as the technology’s reliability when being used. Subthemes for this section include (a) users adapted to voice app’s conversational style, and (b) device unreliability.

### Users Adapting to Voice App’s Conversational Style

Most participants found the device set-up and instructions fairly straightforward, but at times struggled to successfully log their measurements on the *Medly* voice app. When participants struggled, they adjusted the way they spoke instead of continuing in their natural manner in hopes that the voice app would understand them better.

> “I learned how to get into her rhythm as opposed to her getting into my rhythm.” [Participant 04]

Specific strategies were employed to change their speaking style and most often involved modifying the volume, tone, pace, and style they spoke at. Different strategies seemed to work better for different participants, specifically with the pace at which they spoke at.

> “Now I just say 116.4 pounds (faster) and there’s absolutely no issues with her now.” [Participant 12]

> “Of course I would either make sure to be speaking directly at it or elevate my voice or something like that.” [Participant 15]

> “I want to record one hundred, but it’s very typical to say ‘a hundred’ and not ‘one hundred’, but I notice it doesn’t pick up on that.” [Participant 17]

Once participants changed their conversational tone when speaking to the voice app, they began to notice difficulties in the interaction since it no longer felt like a natural conversation.

> “It’s like when you talk to someone foreign or you know from another country or another language and you try to say a few words for them to understand it.” [Participant 12]

> “I try to, like, separate each word, almost like I had to speak robotic.” [Participant 18]

> “I have to be serious, slow and sure of how I say the numbers.” [Participant 17]

Another interaction strategy employed by most participants involved using the device’s touchscreen capabilities. In most cases this alternative input was the favorable approach over using voice since it was simpler to use and most importantly, faster.

> “I got into a routine which allowed me to go through it as quickly as possible, and that routine would be that I would speak the results for weight, blood pressure and heart rate, and then I would interact directly on the touch screen for symptoms so we didn’t have to wait for her. So yes, every time I use the touch screen it works fine and the fact that I could use a touch screen and it would work even though she hadn’t finished speaking is a big plus for me.” [Participant 15]

Interactions were found to be most successful when participants did not multitask on other items.

> “You can multitask if you really want to, but that’s what I think mistakes can be made easier.” [Participant 02]

> “I knew the questions that were going to be asked after a while, but I still listened. Only because you know I’d rather do it right than wrong if I can.” [Participant 08]

Despite the learning curve experienced by most participants, the mitigation strategies described above support the feasibility of deploying a voice app, such as *Medly*, in the clinic due to the perseverance displayed by these participants to make the interaction easier for themselves as time went on.

### Device Unreliability

Almost all participants experienced some level of difficulty when they interacted with the voice app. Sometimes the voice app froze and the session ended abruptly, and other times it would not provide the user with an opportunity to correct any of the wrong measurements.

> “You can go back and correct it, right, but sometimes it gives you a little bit of a hassle so I have to start over.” [Participant 02]

> “Then she just shut down … When she couldn’t get the measurements or something, she would just turn off.” [Participant 04]

Participants also described instances where the voice app was not able to correctly pick up the information they were saying, making them feel frustrated, annoyed, panicked, and discouraged to the point where they no longer wanted to use the device that day.

> “Yeah, I’d wake up in a great mood and oftentimes it was so frustrating that it made me cranky afterwards. Yeah, it really switched my mood. One time she repeated it to me and I thought she got it alright and then she repeated it and said that I fainted and I had not fainted, so I panicked.” [Participant 18]

When the voice app was not able to pick up the correct measurements, participants often felt the need to speak louder. This was considered to be specifically problematic in situations where a participant may not be feeling well and does not have the ability to project their voice. As explained by one participant, with the smartphone they are able to share information without needing to exert a lot of energy:

> “I would never want it to not be on my phone when I go into the hospital and I have a hard time talking. If my blood pressure is through the roof or it’s way too low from retaining water, it’s so hard to speak and I love that I could just throw my phone at the doctor and be like “look, this was [my data] two days ago.” … I really like that.” [Participant 18]

Although the voice app seems feasible to deploy from a patient interaction perspective, users also experienced difficulties when interacting with the device for various tech-related reasons. Understanding the causes and frequencies of these malfunctions will help identify when and where it is appropriate to use voice apps like *Medly*.

### Voice App Acceptability

This theme describes the extent to which the study participants found the *Medly* voice app satisfactory. This level of acceptability includes not only the participant’s thoughts, but also other factors that may influence their experience as described by the following subthemes: (1) device integrated well within household and user lives, (2) users blamed themselves when problems arose with the voice app, (3) voice app missing specific features desired by users.

### Device Integration in Household

In addition to using the device to access the *Medly* voice app, many participants also found they used it for other things during their time on the study. Over the four weeks some participants described the device as a companion, with one participant noting:

> “She became like a buddy. I know it’s little quirks, specifically when it makes mistakes… I would say for people that live on their own or whatever it can become like a friend, right?” [Participant 08]

Some participants also described their experience as “pleasant” when interacting with the device, and others specifically feeling the need to use manners and to be polite while conversing with it:

> “And I’ve gotten along with Alexa just fine. It was so cute. I was inputting on *Medly* and I did it with Alexa at the same time and at the end I said ‘Alexa, thank you’ and she said ‘you bet’… One night I said, ‘oh Alexa goodnight’ and she said ‘night night, sleep well’.” [Participant 08]

Not only did the device become a companion for the user, but for other family members and friends as well:

> “She did give my granddaughter a knock knock joke the other night. [The grandkids] have fun with her by asking what the weather is or something like that.” [Participant 10]

This interaction is an example of how easily the device can fit in and become integrated within a space in the household. While in common areas, users have noted using the device for other activities, such as:

> “I let it play music for me or I ask what’s the weather like today and I do the CTV News first thing in the morning, so yeah, I think it’s a great thing.” [Participant 02]

Having the device in common spaces also served as a reminder for some participants who have difficulty remembering to perform their *Medly* measurements. Others also mentioned that because the device was sitting in a common space, they would be more inclined to use *Medly* on it:

> “Seeing the monitor right there on the counter I feel like it definitely encourages and motivates me and is a visual reminder as opposed to the app on the phone to actually do it.” [Participant 11]

> “At first I thought it would be my phone. But probably you know, now it’s Alexa. She sits right there, so probably Alexa.” [Participant 02]

Some participants also situated the device in other places in their house, such as the bedroom. In these cases they also found the set-up to be useful:

> “I use it at night time when I’m going to bed like you know, relaxing music.” [Participant 06]

And even in some cases, more preferred when compared to the smartphone:

> “I’m in my bedroom and I have a bathroom in the room, so when I go in the bathroom to weigh myself, I do my blood pressure at the same time. So ideally that is where I talk to [Alexa]… over the last week it’s been working and I really like that because then I’m done and then I can go right back to bed after I take my pills so it doesn’t make my mind wake up.” [Participant 03]

> “I’m sort of having concussion symptoms and the phone makes me nauseous. So at the moment, I prefer only having to do it with Alexa.” [Participant 14]

In addition to the benefits of the device integrating well within different spaces in the household, there are also drawbacks that can exist when keeping the device in a public space. Most participants noted the importance of having a quiet space to focus and successfully submit readings:

> “Honestly like I did it more often when I didn’t have my son because everything here he likes to speak over me… He would repeat ‘Alexa’ behind me.” [Participant 18]

> “Like if my husband would walk into the kitchen as I was doing it, I would shoo him away, literally.” [Participant 08]

### Users Blamed Themselves When Voice App Problems Arose

Although frustration was experienced by some participants when the device abruptly stopped working or incorrectly heard them, often times (especially in the first week) users felt like it was their fault when a mistake was made:

> “I wasn’t annoyed by it. I just thought, oh, I’m not speaking clearly or loudly enough, or you know.” [Participant 08]

> “Well again, I go back to the learning curve in the first week. There was some frustration, but you can’t blame that on Alexa, that was all me.” [Participant 05]

These reflections indicate that users were generally understanding of the voice app and had some patience when interacting with it.

### Missing, but Desired Voice App Features

Participants shared some of the features they value in devices that can offer programs such as *Medly* on. In particular, users would prefer to interact with a device that is fast and can quickly record their data for the day. In some instances users compared the voice app to Bluetooth capability, indicating the latter is a much faster and more simpler process:

> “It’s just really cumbersome, like the whole process. And I guess part of that is because the [smartphone] app is so easy. And I think it could get even easier if I got the Bluetooth blood pressure and scale.” [Participant 04]

> “To me, honestly, because they want it in the morning, the smartphone is much faster.” [Participant 09]

Most users also expressed concern about how they would use the voice app should they go on an overnight trip. A device that is small enough in size to be portable when traveling was desired and often mentioned.

> “The only thing I don’t like about it is it is big and bulky so it is not something I would be too inclined to want to travel with. So yeah, so for me the mobility issue would be a bit of a concern if I had to rely on it.” [Participant 13]

## Discussion

### Principal Findings

This manuscript presents the findings from a clinical pilot study for a voice app, designed for HF patients, using a mixed methods approach. To our knowledge, this has been the first clinical pilot evaluation of a voice app used for helping patients manage an advanced chronic condition at home. So far, studies have only reported on accuracy and acceptability levels in a controlled lab environment, however these findings are still consistent with the results presented in this paper (15,16). This study indicates the level of acceptability and feasibility of a voice app for patient self-management by measuring engagement levels. Our findings show that there was a 14% decline in engagement levels between week one and week four levels. Despite engagement decreasing as the weeks progressed, participants became more accepting of the technology as time went on (higher SUS scores when compared to week two). The workload associated with using the voice app was not seen as problematic based on NASA-TLX scores, although participants reported needing to use a higher cognitive load in week four when compared to week two data (4% increase). Regardless of these positive results however, there were 13% more participants who stated they were less interested in using in the future when compared to week two results.

Aside from understanding the acceptability and feasibility of using the voice app as an alternative input for chronic disease management, we also sought to identify who this technology would be best suited for. Similar to the findings presented by Ware et al., engagement levels were highest in the older age group demographic, and progressively lower in younger age groups (17). While the oldest group had the highest engagement levels, the middle-aged demographic (41-60 year old’s) had the highest SUS average score, indicating they were the most accepting of the voice app.

One of the most common responses provided by participants during the interview was the notion that the voice app takes a long time to complete, and in particular, takes longer than the *Medly* smartphone app. Users would often describe being rushed out the door in the mornings, in which case they appreciated being able to use the smartphone app to quickly input their measurements. This type of lifestyle and response was observed less with the older demographics who generally seemed to have more patience and understanding when interacting with the voice app. There were also specific cases where the voice app actually proved to be more useful than the smartphone. One participant was experiencing concussion-type symptoms, and as a result had limited screen time, so the voice app worked well for them. Another participant often felt fatigue as one of the side effects from their medications and experienced difficulties navigating the *Medly* smartphone app in the mornings. In this case, they also appreciated how much easier it was to perform the required tasks using the *Medly* voice app. Similar sentiments were echoed by other participants who came to the realization that they can successfully record their readings when speaking in a relaxed, non-strenuous manner. While this worked well for some, one participant in a similar situation had a different experience, specifically because the voice app was unable to decipher their speech when they were feeling unwell due to their weak and fragile voice. As a result, further advancements are required to better recognize sound, specifically when users are not able to exert large amounts of energy while speaking. Similar technical limitations have also been outlined by other voice app studies (18).

The findings from this study also showcase how well integrated the device became in many households and the potential benefits this may have for participants. Because of the device’s versatility, it quickly became a part of many users’ daily routine, from listening to music to asking for dinner recipes, and started turning into a companion. Not only did the device provide social support, but it also served as a visual reminder to perform their *Medly* measurements. One participant also noted they would be more inclined to use the *Medly* voice app simply because it was in a common space they frequent in their house. Therefore, the natural integration of the device into users’ lives over the four weeks shows the possibility that it may make it more convenient for some to perform their *Medly* measurements, and may encourage and motivate others who often forget.

These findings help begin to uncover the “profile” of the patient demographic this technology would be most suitable for. Data from the clinical pilot show that those who feel more confident in using technology, have less busy schedules, as well as those who are older (60+ years) have an easier time, are more successful and consistent when interacting with the voice app. Also, those with multimorbidity can benefit from using this platform especially due to common side effects they may experience from their conditions.

## Limitations

Multiple limitations were identified over the course of the study and as a result should be acknowledged to better understand the impact of the findings.

First, because there were numerous questionnaires and interviews, the study team was mindful of the potential for social desirability bias (19). As a result, participants were encouraged to speak honestly and were given the opportunity to disclose their thoughts through questionnaires instead of over the phone. Second, our aim was to recruit 30 participants but only 20 patients were onboarded. Therefore, most of the findings and results were interpreted in a qualitative manner since they were not statistically powered. Third, specific study factors could have impacted the participant’s thoughts, experiences, and feedback. Users were aware that the study duration was only a four week period and as a result, may have had higher engagement levels than if they were asked to use it for a longer period of time. Participants were also required to perform a double entry of their measurements, which may have impacted some users’ routines had they only been required to use the voice app. Fourth, because the inclusion criteria was general enough to include any patient enrolled in the program, selection bias likely occurred during recruitment. In this case, there may have been missed opportunities to include a greater variety of demographics in the study, especially those who primarily spoke languages other than English. Lastly, because most participants from this study never interacted with a smart speaker before, their thoughts and feedback may be influenced by the fact that they were interacting with a novel technology. As a result, their thoughts on the device itself could be reflected in their responses, even though any voice user interface device could have been used for the study.

## Conclusions

This study utilized a mixed methods approach to investigate the acceptability and feasibility of deploying a voice app for DTx used in chronic disease management. Our findings were consistent with previous research when it came to engagement levels, with the oldest age group showcasing the best, most consistent results. We recommend this platform be offered to those who: are older (60+ years), have less busy schedules, exhibit high confidence levels when using technology, or experience symptoms (such as fatigue or headaches) from chronic conditions. While the technology could benefit from some advancements, participants were successful in finding ways to improve their conversational experience, proving that an app like this could be feasible to deploy in the clinic for future use.

## Data Availability

The work that was produced and collected in this study is stored in University Health Network (UHN) servers. Our agreement with the UHN Research Ethics Board describes very specific circumstances that does not allow us to make the data publicly available and prohibits us from sharing the data on a public platform.

## Acknowledgments

First, the authors wish to thank the patients who participated in this study. Thank you to the *Medly* nurse coordinators: Mary O’Sullivan, Sarvatit Bhatt, Eva Pavic, Tina Carriere, and Annabelle Fontanilla for helping with the recruitment process. We also wish to express our gratitude to Quynh Pham and Patrick Ware in helping guide this research project’s methodology, as well as Cait Nuun, Madison Taylor, and Denise Ng for their REB expertise and guidance.

## Abbreviations

HF: heart failure
SUS: System Usability Scale
UHN: University Health Network
UTAUT2: Unified Theory of Acceptance and Use of Technology
VUI: voice user interface

## Supporting Information

Multimedia Appendix 1 **Fig. S1 Instructions manual for *Medly* voice app**.

Multimedia Appendix 2 **Table S1. Baseline questionnaire for participants**.

Multimedia Appendix 3 **Table S1. Overall and weekly average engagement levels based on various patient characteristics**.

Multimedia Appendix 4 **Fig S1. SUS score distributions**.

Multimedia Appendix 4 **Fig S2. NASA-TLX score distributions from week two and four results (top and bottom, respectively)**.

Multimedia Appendix 4 **Table S1. Average scores for each NASA-TLX question**.

Multimedia Appendix 4 **Table S2. Average scores for each of the constructs from the UTAUT2 questionnaire**.

## References

1. Noncommunicable diseases [Internet]. World Health Organization. 2021. Available from: https://www.who.int/news-room/fact-sheets/detail/noncommunicable-diseases

2. Dickson VV, Clark RA, Rabelo-Silva ER, Buck HG. Self-Care and Chronic Disease. Nurs Res Pract [Internet]. 2013; Available from: http://dx.doi.org/10.1155/2013/827409

3. Seto E, Leonard KJ, Cafazzo JA, Masino C, Barnsley J, Ross HJ. Self-care and quality of life of heart failure patients at a multidisciplinary heart function clinic. J Cardiovasc Nurs [Internet]. 2011; Available from: http://dx.doi.org/10.1097/JCN.0b013e31820612b8

4. Ware P, Ross HJ, Cafazzo JA, Laporte A, Seto E. Implementation and evaluation of a smartphone-based telemonitoring program for patients with heart failure: Mixed-methods study protocol. J Med Internet Res [Internet]. 2018; Available from: http://dx.doi.org/10.2196/resprot.9911

5. Irfan Khan A, Gill A, Cott C, Hans PK, Steele Gray C. mHealth Tools for the Self-Management of Patients With Multimorbidity in Primary Care Settings: Pilot Study to Explore User Experience. JMIR Mhealth Uhealth [Internet]. 2018 Aug 28;6(8):e171. Available from: http://dx.doi.org/10.2196/mhealth.8593

6. Mahmood A, Kedia S, Wyant DK, Ahn S, Bhuyan SS. Use of mobile health applications for health-promoting behavior among individuals with chronic medical conditions. Digit Health [Internet]. 2019 Jan;5:2055207619882181. Available from: http://dx.doi.org/10.1177/2055207619882181

7. Brownstein J, Lannon J, Lindenauer S. 37 Startups building voice applications for healthcare [Internet]. MobiHealthNews. Available from: https://www.mobihealthnews.com/content/37-startups-building-voice-applications-healthcare

8. Kannel WB, Belanger AJ. Epidemiology of heart failure. Am Heart J [Internet]. 1991 Mar;121(3 Pt 1):951–7. Available from: http://dx.doi.org/10.1016/0002-8703(91)90225-7

9. Ware P, Ross HJ, Cafazzo JA, Boodoo C, Munnery M, Seto E. Outcomes of a heart failure telemonitoring program implemented as the standard of care in an outpatient heart function clinic: Pretest-posttest pragmatic study. J Med Internet Res [Internet]. 2020; Available from: http://dx.doi.org/10.2196/16538

10. Johanson GA, Brooks GP. Initial Scale Development: Sample Size for Pilot Studies. Educ Psychol Meas [Internet]. 2010 Jun 1;70(3):394–400. Available from: https://doi.org/10.1177/0013164409355692

11. Proctor E, Silmere H, Raghavan R, Hovmand P, Aarons G, Bunger A, et al. Outcomes for implementation research: Conceptual distinctions, measurement challenges, and research agenda. Administration and Policy in Mental Health and Mental Health Services Research [Internet]. 2011;38(2):65–76. Available from: http://dx.doi.org/10.1007/s10488-010-0319-7

12. Venkatesh V, Thong JYL, Xu X. Consumer acceptance and use of information technology: Extending the unified theory of acceptance and use of technology. MIS Quarterly: Management Information Systems [Internet]. 2012; Available from: http://dx.doi.org/10.2307/41410412

13. System Usability Scale (SUS) [Internet].usability.gov. Available from: https://www.usability.gov/how-to-and-tools/methods/system-usability-scale.html

14. Nasa. NASA Task Load Index. Human mental workload [Internet]. 2006;1(6):21–21. Available from: http://www.ncbi.nlm.nih.gov/entrez/query.fcgi?cmd=Retrieve&db=PubMed&dopt=Citation&list_uids=16243365

15. Jadczyk T, Kiwic O, Khandwalla RM, Grabowski K, Rudawski S, Magaczewski P, et al. Feasibility of a voice-enabled automated platform for medical data collection: CardioCube. Int J Med Inform [Internet]. 2019;129(March):388–93. Available from: http://dx.doi.org/10.1016/j.ijmedinf.2019.07.001

16. Cheng A, Raghavaraju V, Kanugo J, Handrianto YP, Shang Y. Development and evaluation of a healthy coping voice interface application using the Google home for elderly patients with type 2 diabetes. In: 2018 15th IEEE Annual Consumer Communications & Networking Conference (CCNC) [Internet]. IEEE; 2018. Available from: http://dx.doi.org/10.1109/ccnc.2018.8319283

17. Ware P, Dorai M, Ross HJ, Cafazzo JA, Laporte A, Boodoo C, et al. Patient Adherence to a Mobile Phone–Based Heart Failure Telemonitoring Program: A Longitudinal Mixed-Methods Study (Preprint) [Internet]. Available from: http://dx.doi.org/10.2196/preprints.13259

18. Sezgin E, Noritz G, Elek A, Conkol K, Rust S, Bailey M, et al. Capturing at-home health and care information for children with medical complexity using voice interactive technologies: Multi-stakeholder viewpoint. J Med Internet Res [Internet]. 2020; Available from: http://dx.doi.org/10.2196/14202

19. Grimm P. Social Desirability Bias. In: Wiley International Encyclopedia of Marketing [Internet]. 2010. Available from: http://dx.doi.org/10.1002/9781444316568.wiem02057

